# Effectiveness of gamified team competition in the context of mHealth intervention for medical interns: a micro-randomized trial

**DOI:** 10.1101/2022.03.11.22272278

**Authors:** Jitao Wang, Yu Fang, Elena Frank, Maureen A Walton, Margit Burmeister, Ambuj Tewari, Walter Dempsey, Timothy NeCamp, Srijan Sen, Zhenke Wu

## Abstract

**Background:** Twin revolutions in wearable technologies and smartphone-delivered digital health interventions have significantly expanded the accessibility and uptake of personalized interventions in multiple domains of health sciences. Gamification, the application of gaming elements to increase enjoyment and engagement, has the potential to improve the effectiveness of digital health interventions. However, the effectiveness of competition gamification components remains poorly understood, challenging informed decisions on the potential adoption of these components in future studies and trial designs. We aimed to evaluate the effect of smartphone-based gamified team competition intervention on daily step count and sleep duration via a micro-randomized trial.

**Methods:** We recruited first-year medical residents (interns) in the US, who downloaded the study app, provided consent, wore a wearable device, and completed a baseline survey. Teams were formed based on participating residents’ institutions and specialties, and subsequently randomized weekly to the competition or non-competition arms. In the competition arm, opponent teams and competition type (step count or sleep duration) were also randomly selected. Competition participants had access to the current competition scoreboard and competition history via the study app; they also received scheduled competition-related push notifications in a competition week. We estimated the main and moderated causal effects of competition on proximal daily step count and sleep duration. This trial is registered with ClinicalTrials.gov (NCT05106439).

**Findings:** Between April and June 2020, we enrolled 2,286 medical interns from 263 institutions, of whom 1,936 were formed into 191 teams that met the criteria for participation in competitions between July 6 and September 27, 2020. 1,797 participants who had pre-internship baseline information were included in the analysis. Relative to the no competition arm, competition intervention significantly increased the mean daily step count by 111·5 steps (SE 40·4, p=0·01), while competition did not significantly affect the mean daily sleep minutes (p=0·69). Secondary moderator analyses indicated that, for each additional week-in-study, the causal effects of competition on daily step count and sleep minutes decreased by 9·1 (11·6) steps (p=0·43) and 1·9 (0·6) minutes (p=0·003), respectively. Intra-institutional competition negatively moderated the causal effect of competition upon daily step count by −114.9 (93·7) steps (p=0·22).

**Interpretation:** Gamified competition delivered via mobile app significantly increased daily physical activity which suggests that team competition can function as a mobile health intervention tool to increase short-term physical activity level.

**Research in context:** *Evidence before this study:* We searched PubMed for studies of mobile health intervention with gamified components: (“mobile health intervention”, “mHealth intervention”, “mobile health gamification”). We evaluated studies published before November 30, 2021. The search was not limited by language. Previous work affirmed that in mobile health interventions, gamification is effective for improving user’s physical activity and mental health. Most of previous work used feedback, reward, and progress bar as game mechanics, while none have rigorously examined the effectiveness of gamified team competition.

*Added value of this study:* This study provides evidence that the gamified team competition has a positive effect on physical activity. The data that was intensively collected as part of this study can be used for further investigation.

*Implications of all the available evidence:* The results of this study indicate that gamified team competition has the potential to improve the effectiveness of and engagement with mobile health interventions.

## Introduction

Sufficient physical activity and sleep are associated with lower risk for numerous health conditions, including cardiovascular disease, obesity and depression^1–3^. However, only one in four US adults meets the recommended 150 minutes of moderate-intensity activity per week^4^, and over one-third of US adults do not achieve the recommended seven hours of sleep per night^5,6^.

Recent technical advances in wearable devices and mobile phones provide a new integrated platform to deliver interventions with minimal expense and user burden^7^ with the additional advantage of temporal and spatial flexibility. Mobile devices can collect real-time and objective measurements of a user’s physical activity and geographic location to provide personalized just-in-time adaptive interventions (JITAI)^8^. To date, many previous studies have shown the effectiveness of wearable and smartphone-based intervention on health outcomes^9–11^. Some studies included gamification, a strategy that attempts to enhance user enjoyment and engagement^12^ by introducing game mechanics into a non-game environment^13,14^. Theories of health behavior change suggest that gamification elements that prompt self-monitoring, such as performance feedback, progress monitoring, and social comparison have the potential to motivate changes in behavioral outcomes^15,16^.

Team competition is one such potential gamification strategy, however to our knowledge, its effectiveness at improving health behaviors has not been formally assessed.

Micro-randomized trials (MRT) can be used to address scientific questions about whether and under what circumstances JITAI components are effective, with the ultimate goal of developing effective and efficient JITAI^17–19^. In this study we conducted an MRT using principles of health behavior change and gamification to deliver a mobile app-based weekly team competition to evaluate the effectiveness of this type of mHealth intervention on individual physical activity, sleep duration in the population of medical intern. We also explore the effectiveness of this mHealth intervention on user’s engagement and individual self-reported mood score, inspired by previous work showing that increased sleep opportunity and physical activity may improve individual’s mood in this depression-vulnerable population^20,21^. Although we mainly focused on the effect of team competition on short-term (proximal) outcomes including step count and sleep minutes in this study, our ultimate goal is to improve interns’ long-term (distal) mental health by increasing their short-term physical activity and sleep duration.

The study was conducted among a national cohort of first-year medical residents. Medical internship, a one-year-long physician training program, is highly stressful, which may lead to reduced health functioning and mental health symptoms. This study also assessed potential effect moderators, variables that increase or decrease the effectiveness of mHealth intervention to inform future research incorporating personalized team-competition into mHealth intervention.

## Methods

### Study design and participants

We conducted a three-month MRT to investigate the causal effects of team competition upon proximal weekly average daily step counts, minutes of sleep, participation rate and mood score via the Intern+ mobile app as part of the Intern Health Study, a prospective cohort study assessing stress and depression during the first year of residency training in the USA^22^. Training physicians, who began their internship in July 2020, were invited via email to participate in the study within one to three months prior to the start of internship. Ownership of an iPhone supporting iOS 10·0 or later or an Android device supporting version 6·0 or later was required. Upon enrollment, participants were provided with a Fitbit Charge 3 to collect sleep and activity data if they did not already own a compatible Fitbit or Apple Watch. All participants provided informed consent electronically and were compensated $80 to $130. The University of Michigan institutional review board approved the study.

To protect participant anonymity, we required a minimum of five participating interns per team to be eligible for the competition arm of the study. Programs with at least five interns were grouped into program-based teams (e.g., “Michigan Psychiatry”). Interns within the same residency institution in programs that did not meet this criterion were grouped into institution-based teams (e.g., “Michigan Programs”), also with a minimum of five participants per team. All the remaining enrolled study subjects were considered ineligible for the competition arm.

All the eligible subjects were onboarded before their internships started on July 1, 2020. Baseline surveys that assessed interns’ stress; also, baseline step counts and sleep minutes were recorded via the Fitbits/Apple Watch. The competition assignment started on the first Monday following the start of internship (July 6, 2020) and ended on Sep 27, 2020 (Sunday of the 12th week), which lasted nearly three months. Each competition episode was one week, starting on Monday 00:00 and ended on Sunday 23:59.

### Randomisation and masking

Each week, we repeatedly randomized interns by three factors: competition status (in competition or not), opponent team, and competition type (on step count or sleep minutes). Such a factorial design enables inference of causal effects of one or multiple factors based on the same study data. In particular, first, each team was randomized with equal probabilities to the competition or non-competition arm every Monday - this is the main randomization of the study. Second, every week, teams in competition were randomly assigned an opponent team: 1) total randomization, where the opponent team was assigned regardless of institution and specialty (e.g., Michigan Pediatrics vs Yale Emergency Medicine); 2) intra-institutional randomization, where two competing teams were from the same institution (e.g., NYU Internal Medicine vs NYU Surgery); 3) intra-specialty randomization, where two opponent teams were from the same specialty (e.g., Northwestern Psychiatry vs OSU Psychiatry). All the three rules for opponent assignment had equal probability (1/3) to be selected for each week. If there were an odd number of teams inside the randomization pool, then the team left over would be put back into the non-competition arm. Third, for each pair of opponent teams, there was a 50/50 chance of competing on average daily step counts or average daily sleep minutes. Figure 2 details the randomization scheme. Due to the nature of the intervention, participants could not be masked from the competition assignment. Although investigators were not masked to intervention allocation, all data collected from participants was through the app or wearable device.

**Figure 1:**
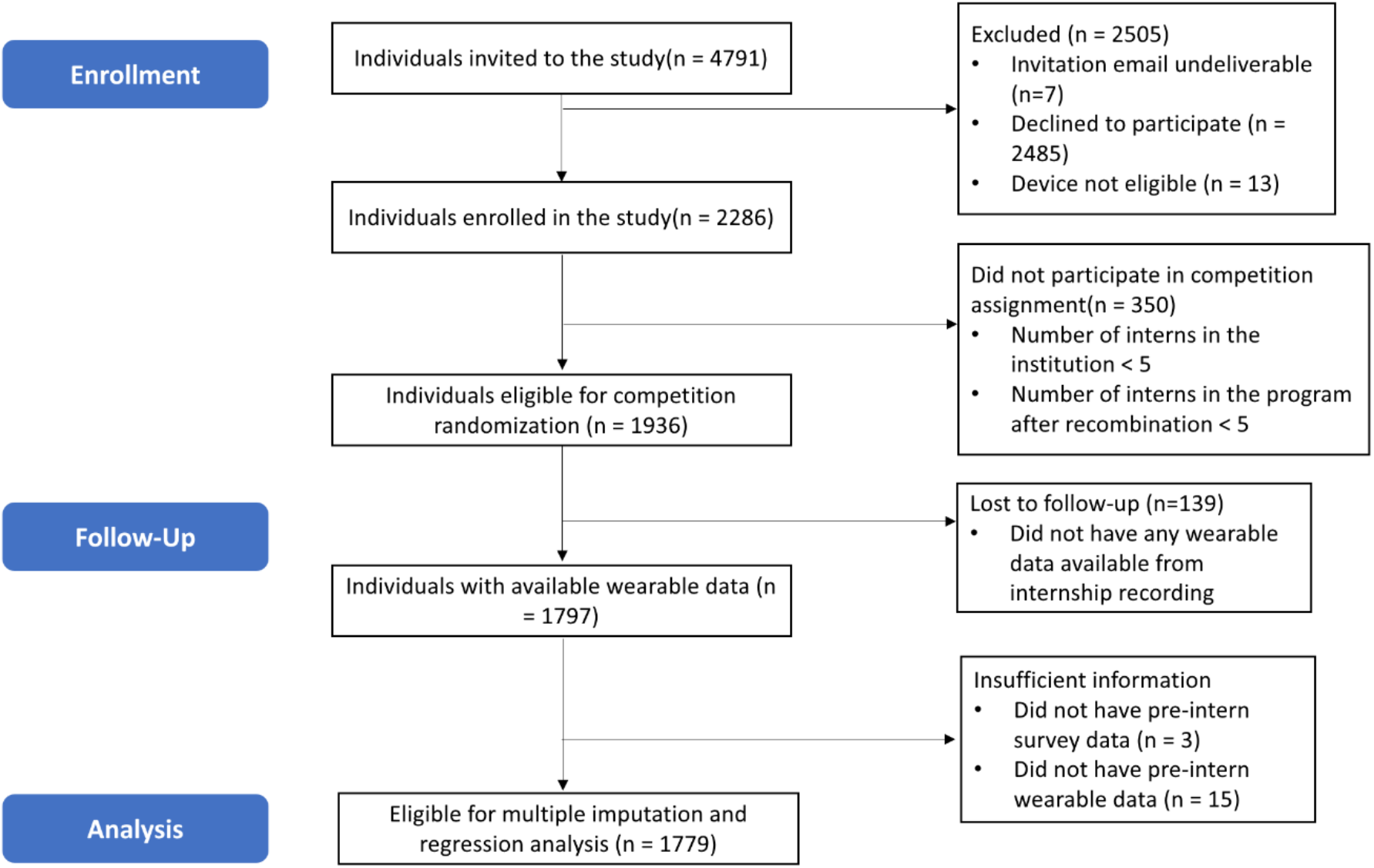
Flow diagram detailing subject inclusion from enrollment through analysis.

**Figure 2:**
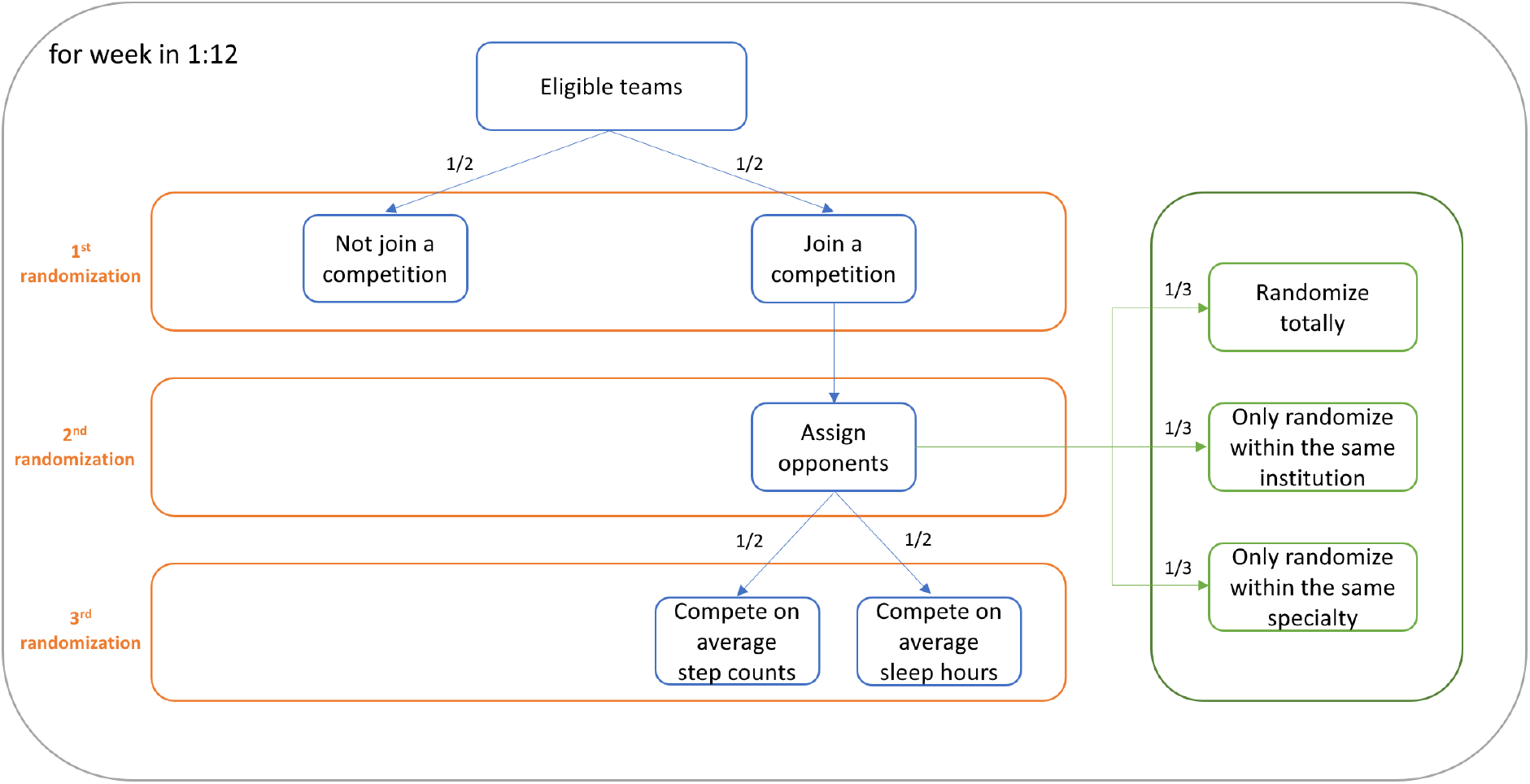
Randomization scheme of Intern Health MRT.

### Procedures

After completing consent and downloading the study app, the wearable devices started to record daily step count and time spent asleep. Participants were prompted to report their daily mood (a score of 1 corresponded to the lowest and a score of 10 corresponded to the highest mood) every day at a user-specified time between 5 PM and 10 PM (default was 8PM) in the study app. In addition to collecting data, the study app aggregated and displayed visual summaries of participant’s historical data, including daily step count, sleep minutes and mood score, through a dashboard which participants could access at any time via the app (eFigure 1). Separate from the competition component of the app, each user also had a 50/50 chance each day to receive a push notification at 3pm which contained a message summarizing their personalized data feedback, a relevant fact or tip for improving mental health and well-being, or a general supportive statement. Furthermore, baseline and quarterly follow-up surveys were administered through the app.

The competition intervention was conducted for 12 weeks (Monday July 6, 2020, to Sunday September 27, 2020). Each team was randomly assigned to competition (intervention) and non-competition arm during each competition episode. Four competition-related smartphone push notifications were sent to participants in each competition week:1) an alert of competition type (steps or sleep) and opponent team (Sunday 9 pm prior to the competition week), 2) two competition score updates (Wednesday 9 pm and Saturday 11 am during the competition week), and 3) the final competition results (Monday 12 pm following the competition week). Examples of messages are included in eTable 1. Participants could view their current competition scoreboard and competition history at any time via the Intern+ app. eFigure 1 shows three representative screenshots of the app interface involving competition.

**Table 1:** Demographics characteristics and specialty for study participants (N=1779)

| Demographic characteristics |  |  | Specialty | N (%) |
| --- | --- | --- | --- | --- |
| Age (years), mean (SD) | 27·6 (2·6) |  | Internal medicine | 492 (27·7) |
| Sex (Female), N (%) | 969 (54·5) |  | Surgery | 240 (13·5) |
| Pre-internship baseline steps/day, mean (SD) | 8121·0 (3228·9) |  | Pediatrics | 215 (12·1) |
| Pre-internship baseline sleep mins/day, mean (SD) | 420·6 (107·5) |  | Emergency medicine | 145 (8·2) |
| Internship average days of competition, mean (SD) | 40·7 (13·5) |  | Psychiatry | 126 (7·1) |
| Race, N (%) |  |  | Ob/Gyn | 104 (5·8) |
| *White* | 957 (53·8) |  | Anesthesiology | 94 (5·3) |
| *Black/African American* | 106 (6·0) |  | Family medicine | 78 (4·4) |
| *Hispanic/Latino* | 96 (5·4) |  | Neurology | 49 (2·8) |
| *Asian* | 420 (23·6) |  | Med/Peds | 38 (2·1) |
| *Arab/Middle Eastern* | 31 (1·7) |  | Transitional | 21(1·2) |
| *Other/Multiracial/Not reported* | 169 (9·4) |  | Other | 177 (9·9) |

**Table 2:**
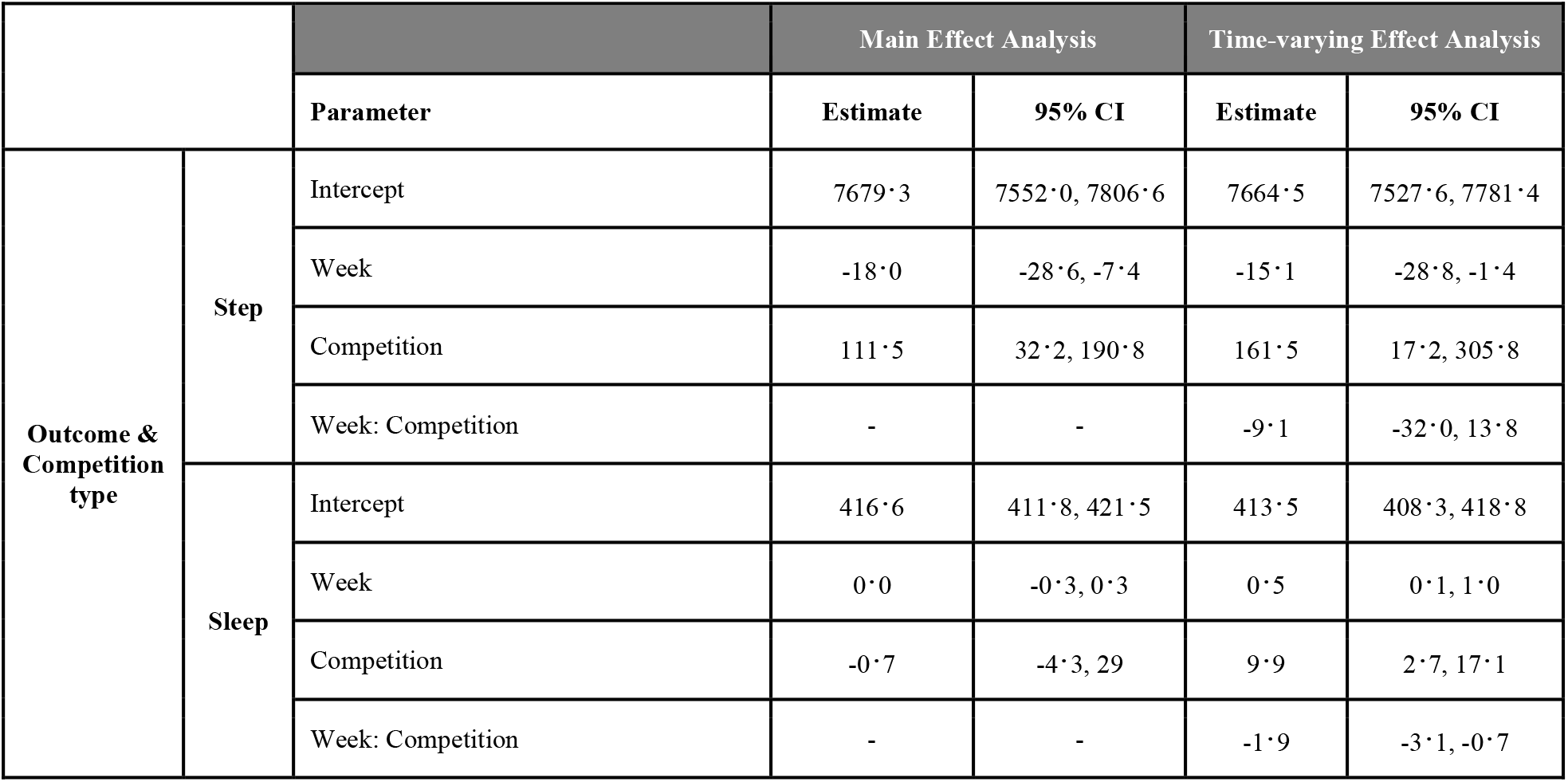
Parameter estimates for linear assessing marginal and time-varying causal effect competition on daily step count and sleep duration.

|  |  |  | Main Effect Analysis |  | Time-varying Effect Analysis |  |
| --- | --- | --- | --- | --- | --- | --- |
|  |  | Parameter | Estimate | 95% CI | Estimate | 95% CI |
| Outcome & Competition type | Step | Intercept | 7679·3 | 7552·0, 7806·6 | 7664·5 | 7527·6, 7781·4 |
|  |  | Week | -18·0 | -28·6, -7·4 | -15·1 | -28·8, -1·4 |
|  |  | Competition | 111·5 | 32·2, 190·8 | 161·5 | 17·2, 305·8 |
|  |  | Week: Competition | - | - | -9·1 | -32·0, 13·8 |
|  | Sleep | Intercept | 416·6 | 411·8, 421·5 | 413·5 | 408·3, 418·8 |
|  |  | Week | 0·0 | -0·3, 0·3 | 0·5 | 0·1, 1·0 |
|  |  | Competition | -0·7 | -4·3, 29 | 9·9 | 2·7, 17·1 |
|  |  | Week: Competition | - | - | -1·9 | -3·1, -0·7 |

### Outcomes

The primary outcomes of the study were proximal weekly average daily step count and sleep minutes, by taking average values of team members within a competition episode, which were measured by wearable devices (Fitbit or Apple Watch). The exploratory outcome was proximal weekly average daily participation rates of step count and sleep minutes, which was defined as the proportion of days that the participants in the team provided daily step/sleep minutes within a competition week. The daily step count or sleep duration would be missing if the user was not wearing wearable devices during the daytime or nighttime. Demographic information and psychology-related scores were collected from baseline surveys (Table 1).

### Statistical Analysis

Note that the competition assignment was randomized on the team level, therefore all the competition-related analyses in this paper were performed on the team level using summary measurements from each team, that is, the team was treated as the unit of analysis instead of individual. Weekly team-based summary measurements were calculated by taking the average of individuals’ measurements within each team.

The primary aim of this study assesses whether there was a main causal effect of being in the competition arm on the team’s average proximal weekly average daily step count and sleep minutes, compared to not being in the competition arm. The primary analysis was done by fitting linear regression models using generalized estimating equation with independent working correlation matrix (R version 4·0·2; *geeglm* function from R library *geepack*) for average daily step count and sleep minutes separately, with competition assignment, number of weeks in study and control variables. Daily step count and sleep minutes were treated as continuous variables. The competition assignment variable was binary, with a value of 1 for being in a competition week and 0 for a non-competition week. The week-in-study was a continuous variable, with 0 for the first week of the study and 11 for the last week. Percentage of female, team average pre-intern measures, including daily step count, sleep minutes, psychology-related scores (e.g., PHQ-9 score), as well as team average previous week’s outcomes (i.e., previous week’s step count will be included if the outcome is current week’s step count), were included as control variables to increase the statistical power. The procedure implements a weighted and centered least square estimator (WCLS, details included in eMethods), proposed by Boruvka et al^18^.

The secondary aims included moderation analyses to assess potential time-varying effect moderators, aimed at informing the design of real-time personalized and optimized delivery of mHealth intervention. Time-varying effect moderators are moderators that can change the treatment effect and are time-varying because the values of moderators can vary across time (e.g. week-in-study and opponent team)^17^. Two potential time-varying moderators of causal effect of competition were examined. The first moderation analysis is motivated by the hypothesis that the longer a participant was in the study, the more they may be accustomed to the competition intervention or become overburdened, leading them to become less responsive. Interaction terms between number of additional weeks in study and intervention variable were included in the model to evaluate the effect moderation. The second moderation analysis is motivated by the hypothesis that participants in the same institution or specialty tended to have stronger social connection, which may result in fiercer competition to boost the competition effect. Therefore, moderator analysis for whether intern was competing within the same institution or specialty was done by including two additional interaction terms between the intervention indicator and the intra-institution and intra-specialty indicators, respectively.

Exploratory analyses included assessing the causal effects of team competition upon user’s engagement and mood score averaged by team respectively. Linear probability model was used to assess the main causal effect of competition on proximal weekly average daily participation rates of step count and sleep minutes, motivated by the hypothesis that competition intervention can improve user’s engagement to the study app. The casual effect of team competition on team-averaged self-reported mood score was evaluated similarly as our analysis on step count (sleep minutes) in the main and secondary aims.

All the analyses above were based on the weekly aggregated data because every week was treated as a complete episode of competition or not. For the moderation analyses, the results of these effects were reported from the models with linear moderators. Additional results from nonlinear analyses were detailed in eResults.

Multiple imputation was used for individual’s daily step count and sleep minutes to create completed data sets for proper statistical inference so that exactly seven daily values contribute to each weekly average value^23^ (R version 4·0·2; *mice* function from R library *mice*).

## Results

Between April 1, 2020, and June 16, 2020, a total of 4,791 incoming interns received the invitation email and 2,286 (47·7%) of interns enrolled in the study. Of those who enrolled, 84·7% (1,936/2,286) of participants could be grouped in a team with at least five interns and were included in the competition arm, with a total of 191 teams. These eligible competition-arm participants were randomized according to Figure 2. Of the 1,936 participants, 139 (7·2%) participants did not have any fitness tracker data available during the study and 18 (0·9%) did not have sufficient pre-internship survey data and baseline data, which were excluded from the analysis. All remaining interns represented 90 residency institutions and 12 specialties. Among the 1779 (91·9%) participants included in the analysis, the mean age of the participants was 27·6 (SD 2·6), Males and females were nearly equally represented (54·5% female).

Of the 1,779 medical interns who were eligible for intervention, all of them were assigned to the competition arm at least once during the study and the mean number of weeks a participant was in the competition arm was 5·8 (SD 1·9) weeks.

Main-effect analysis indicated that intervention of competing on step count had a significant positive causal effect on proximal daily step count compared to the non-competition arm. The number of daily steps increased by 111·5 (SE 40·4) steps for participants in the competition-step arm, compared to the non-competition arm (p=0·01). While no statistically significant effect on sleep duration was observed in response to competition on sleep. The estimate for the competition-sleep effect is −0·7 (SE 1·8) minutes (p=0·69).

Moderation analyses were performed by adding linear interaction terms between effect moderator and intervention into the model. A negative though non-significant association was observed between the number of weeks in the study and competition-step intervention (−9·1 steps/day; SE 11·6; p=0·43). That is, the moderation analysis (not the main-effect analysis) indicated that being in a competition-step week resulted in about 161·5 additional steps/day during the first week of the study, about 115·8 additional steps/day during the sixth week of the study, and about 61·0 additional steps/day during the twelfth week of the study. Similarly, a significantly negative interaction between the competition-sleep intervention and number of weeks in the study was identified: the causal effect of being in a competition-sleep week changed by −1·9 minutes/day (SE 0·6) with each additional week in the study (p=0·003); note that at the beginning of the study (the first week), the causal effect was significantly positive 9·9 minutes/day (SE 3·6, p=0·008), but then decreased. Plots of estimated causal effects of competition on proximal step count or sleep duration at different weeks and a sensitivity analysis to the linearity assumption (that the causal effect changes linearly by additional weeks in the study) was provided in eFigure 3. We also assessed whether the causal effect of competition upon step count or sleep minutes (relative to no competition) would vary by the opponent team being from the same or a different institution or specialty (eTable 4). Non-significant intra-institution negative moderation (−114·8 steps/day; SE 93·7; p=0·22) and intra-specialty positive moderation (26·1 steps/day; SE 74·7; p=0·73) of causal effect of competition on step were observed. Similarly, no statistically significant intra-institution (0·4 minutes/day; SE 3·2; p=0·90) or intra-specialty moderation (−1·9 minutes/day; SE 3·2; p=0·57) of causal effect of competition on sleep duration were observed.

Exploratory analyses assessed the causal effect of being in a competition week on the proximal daily participation rates of step count and sleep minutes, averaged over all weeks. The positive causal effect on daily participation rates of step count and sleep minutes were a 0·4% (SE 0·3%, p=0·13) and 0·9% (SE 0·3%, p=0·003) respectively. That is, if 1,779 participants were all in the competition week, there would be additional 50 (1,779 * 0·4% * 7) person-day records of step count and 112 (1,779 * 0·9% * 7) person-day records of sleep minutes recorded within this week, compared to a non-competition week. We also assessed whether the causal effect of team competition had a positive impact on team-averaged mood score. Non-significant positive effect (0·02 units/day, SE 0·02, p=0·35) of causal effect of team competition on mood score was observed (eTable 6).

## Discussion

This study answered two questions: 1. Is gamified competition delivered via mobile app effective in the field of mHealth intervention? 2. If it is effective, how to personalize and optimize the efficacy of competition intervention? The main-effect analysis indicated that the gamified competition administered through smartphones can lead to increased proximal daily step count. Positive causal effect suggested inclusion of competition via mobile app is a beneficial component of mHealth intervention. The moderator analysis demonstrated that week-in-study negatively moderates the efficacy of team competition, suggesting a waning causal effect of competition over time. Also, intra-institutional competition decreased the efficacy of competition, suggesting potential improvements in strategies of assigning opponent teams to boost the effect of competition interventions.

The finding on the beneficial effect of mobile-based gamified competition upon physical activity is consistent with previous studies that have shown that the mHealth intervention with gamified components can increase physical activity^24–26^. However, the effect size from our study is smaller than previous studies^25,26^. Under the highly stressful and intensive working environment, the medical interns may be less responsive to the intervention, which may explain smaller effect size relative to other study populations. On the other hand, our result may be applied to other shift workers or under-stress populations who are similar to medical interns.

One possible explanation for the waning causal effect of competition was that interns might be motivated when study began and get tired later so that they were less responsive to the competition assignment. The phenomenon of waning treatment effect is common in the field of mHealth intervention^10,27,28^ and further studies are needed to investigate how to extend the mHealth intervention effect. For example, a break could be given to the interns after an episode of competition assignment to decrease their fatigue, and then intervention can be reintroduced to them after some rest time to regain the benefits from intervention. Adding novel competition-related elements such as levels, scoreboard, and prizes could be another option.

One possible explanation for the negative impact of intra-institutional competition on the causal effect of competition on step count was that perhaps interns felt less competitive within their institution relative to extra-institutional members because they see their fellow institution members as colleagues. The negative moderation of intra-institutional competition suggested avoiding intra-institutional competition assignment in the future application to maximize the competition effect.

We also explored the causal effect of team competition on user’s engagement and mood score. A significant and positive effect of competition on participation rate of sleep minutes was observed, indicating that the competition might have potential to increase user’s engagement. In addition, the positive while non-significant causal effect of competition on mood score was observed.

Our study has multiple strengths. First, compared with standard single-time-point randomized controlled trial design, which can only inform moderation of causal effect by baseline variables (e.g., age, gender), micro-randomized trial enabled us to assess both causal effects of intervention components and time-varying moderation of these effects. Second, a relatively large sample size (1,779 participants in 191 teams) and long study period (12 weeks) allowed us to detect the causal effect of intervention, as well as effect moderators of interest. Third, the unique study population, medical interns with inherent hierarchical structure (by institutions and specialties), allowed us to assess the moderation of social connection and cooperation on the causal effect of gamified competition. Fourth, the analytical approaches we used in the study, the weighted and centered least squares estimator and multiple imputation, allowed us to assess the causal effect moderation consistently and robustly without requiring strong assumptions.

However, there are several unanswered questions that should be addressed in future research. It remains unclear why competition did not affect participant’s sleep duration in the same way as step count. One of our conjectures is that the highly demanding working schedule during medical internship makes interns have little control over their sleep schedule, leading to insensitivity to competition intervention. Also, the reason that intra-institutional competition leads to negative moderation of causal effect of competition needs to be addressed in the future study.

Our study does have several limitations. The first is the data missingness and imputation. More than 30% data were missing for daily step count and 50% for daily sleep duration on individual level. Multiple imputation was used to impute the missing entries under the assumption of missing at random, however, the imputed values of a participant borrowed information from participants of other teams due to limited information in each team, which may result in attenuated estimate when assessing the moderation of competing within the same institution on causal effect of competition since the difference among teams can become smaller after imputation. Second, heterogeneity between Apple Watch and Fitbit charge activity monitors was not accounted for during the analysis. Previous studies have shown that Fitbit Charge 2 and Apple Watch 2 had similar accuracy in terms of estimating step counts^29,30^, however, due to longer battery life, the Fitbit Charge series is more likely to be worn continuously, thus more likely to yield a higher step count and longer sleep duration in real life settings, compared with Apple Watch. Third, the results of this study may not extrapolate to a more general population because medical interns are different from the general population in terms of age, education level, and stress level. Individuals in the general population may be more responsive to the mHealth intervention than medical interns due to more flexible time. Therefore, to validate the generalizability of the results in and out of Intern Health cohort, these suggested interventions should be further refined and replicated in additional studies and cohorts. Fourth, note that instead of individual level analysis, cluster level analysis, where each team was treated as the unit of analysis, was used to avoid ignoring the inherent clustering (team) structure using the team-level summary measures. The summary measures were calculated by taking the average of individuals’ measurements of the same team, which did not account for the heterogeneity among members within the team, resulting in reducing the power of the study. Further methodological research on statistical tools allowing analysis at the level of the individual while accounting for the clustering in the data in the field of MRT is needed.

## Conclusions

In summary, through this smartphone-wearable-based prospective micro-randomized trial, we were able to identify the positive causal effect of competition on proximal step count; exploratory analysis also suggested competition may improve user’s engagement with the study app. In addition, the causal effects of competition were negatively moderated by week-in-study and intra-institutional competition. The competition intervention had no significant causal effect on the sleep duration. These results suggest that gamified competition is worthy of inclusion in the mHealth intervention. Effect of gamified competition may be further boosted by introducing occasional breaks to mitigate waning effects over time and optimizing opponent assignment.

## Data Availability

All data produced in the present study are available upon reasonable request to the authors

## Contributors

All authors made substantial contributions to the study conception and design. S.S. made substantial contributions to the acquisition of data. J.W., Y.F., Z.W. conducted statistical analysis. All authors made substantial contributions to the interpretation of data. J.W., Y.F., E.F., S.S., Z.W. drafted the first version of the manuscript. All authors contributed to critical revisions and approved the final version of the manuscript.

## Data sharing

Deidentified data supporting the results and figures in this manuscript are available upon reasonable request and completion of a data agreement with the Intern Health Study team.

Code for data preprocessing and statistical analysis will be made available online at https://github.com/jtwang95/IHS_competition.

## Acknowledgements

This study was supported by grants from the National Institute of Mental Health (R01 MH101459) to S.S. and Z.W., and an investigator grant from Precision Health Initiative at University of Michigan, Ann Arbor to Z.W. and S.S.. We thank the interns and residency programs who took part in this study.

