## Supplementary appendix for "Effectiveness of gamified team competition in the context of mHealth intervention for medical interns: a micro-randomized trial"

- Page 3: eMethods
- Page 6: eResults
- Page 8: eFigures
- Page 14: eTables
- Page 25: Reference

**eMethods**

**Missing data**

Missing data occurred throughout the trial for various reasons: forgetting to wear a fitness tracker, only wearing fitness tracker during the day, technical glitches, and so on. Therefore, we used multiple imputation, a robust method for dealing with missing data, to impute the daily step count and minutes of sleep. For each day, the daily step count, sleep minutes and mood score were imputed with predictor variables including step count, hours of sleep and mood score from the previous three days, weekly average step count, sleep minutes and mood score from previous week and individual’s baseline characteristics including gender, depressive symptoms score (PHQ-9), neuroticism, early family environment. To accommodate the heterogeneity between different institutions and specialties, individual’s institution and specialty were added to the predictor variable list of imputation. R package ‘mice’ was used to do the multiple imputation and predictive mean matching (pmm) was selected as the imputation method. Results were pooled using 20 imputed dataset following Rubin’s rules.

**Models for assessing causal effect of competition on participation rate**

To investigate the causal effect of competition on intern’s participation rate on daily step count and sleep duration, linear probability model was used to model the mean structure. More specifically, we modeled the probability that the daily step count or sleep duration was measured in week $t$ as $E(Y_{t}|X_{t},W_{t},Z_{t})=\beta_{0}+\beta_{1}Z_{t}+\beta_{2}W_{t}+\beta_{3}X_{t}$ for marginal-effect model, and $E(Y_{t}|X_{t},W_{t},Z_{t})=\beta_{0}+\beta_{1}Z_{t}+\beta_{2}W_{t}+\beta_{3}Z_{t}W_{t}+\beta_{4}X_{t}$for time-varying-effect model, where $Y_{t}$ is a continuous variable between 0 and 1 and the value of $Y_{t}$ represents the proportion of available data points during week $t$ for each team. $Z_{t}$is a binary treatment variable, where $Z_{t}=1$implies a competition week and $Z_{t}=0$ implies a non-competition week. $X_{t}$ is the set of control variables, including team-average baseline data measured before the weekly randomization, for the purpose of reducing variation in the outcome of interest $Y_{t}$. The set of variables $X_{t}$ consists of percentage of female, team-average pre-intern daily step count, sleep minutes and psychology-related scores.

In the Intern Health Study data, whether the intern had a daily step count, sleep duration or self-reported mood survey is considered as a binary outcome. A conventional approach to model the binary outcome is to fit a logistic model. However, in our analysis, we did not adapt conventional logistic regression approach on daily data. Instead, we perform a weekly analysis using linear probability model and use a weighted and centered estimator proposed by Boruvka et al. to estimate the parameters of interest. The reason that we preferred a weekly analysis to a daily analysis is that the competition assignment was randomized on a weekly level and the daily level analysis violates the positivity assumption of the estimating method proposed by Boruvka et al^1^. Besides, the reason that we used linear probability model rather than beta regression model, which is more commonly used than linear probability model for proportional outcomes due to unit interval boundary, was that the theoretical guarantee of using beta regression to assess the time-varying causal effect moderation is not well-established. We calculated the predicted probability for each week in our dataset and all the estimated values fell between unit interval, suggesting that the linear probability model is appropriate in our case.

**Estimation techniques**

To estimate the coefficients of interest with the existence of time-varying moderators, we used a weighted and centered least squares estimator proposed by Boruvka et al^1^. The method uses both the estimating equation method and robust “sandwich” estimate to provide consistent estimates and robust inference. The main advantage of the proposed method is that it does not require correct specifications to the terms that do not interact with treatment, in order to provide consistent estimates for the parameters of interest.

The method proposed by Boruvka et al. can provide valid inference for treatment-related variables when the treatment assignment probabilities are time-varying. Since the treatment assignment probabilities were constantly 0.25 across weeks in the IHS 2020 (each team had 50/50 chance to be in a competition week and for teams in competition week, they had 50/50 chance to compete on step count or sleep minutes), the weights in the estimating equation are all 1. In addition, the centering term for treatment variable is always a constant 0.25.

The estimating equation method with robust error estimation has two main advantages: 1. It does not require distributional assumptions on continuous outcomes. 2. It allows dependence between observations in the data, where the observations for the same team in our dataset are correlated due to repeated measurements.

**Implementation**

Due to the constant weights and centering term in the estimation equation, the estimating problem in our case can be solved by applying standard generalized estimation equation (GEE) method with independent working correlation matrix. We used the standard *geeglm* function in R package *geepack* to obtain the estimates as well as robust standard errors.

Due to the existence of data missingness in the study, multiple imputation with predicted mean matching was used to impute the missing entries. *mice* function in R package *mice* was used to perform multiple imputation and *pool* function in R package *mice* was used to combine the estimates and standard errors from each imputed dataset through Rubin’s rules.

**Mood-score related analysis**

Mental health disorders such as anxiety and depression are considered to be closely related to insufficient physical activity and sleep duration^2,3^. Therefore, we performed analyses on assessing the marginal and time-varying causal effect of competition on intern's mental health outcome: self-report mood score. We also did similar analyses on the participation rate of daily mood surveys. During mood-related analyses, the treatment variable competition was defined as either an intern was in competition step or sleep group. Here, we considered the competition indirectly affected the intern’s mood score since interns were competing on daily step count and sleep minutes, rather than directly on mood score.

**Sensitivity analysis**

To investigate the robustness of the results, we performed three types of sensitivity analysis.

First, we compared the results from complete-case analysis and multiple imputation to evaluate the sensitivity of missing mechanisms.

Second, we used a linear model for the moderation in the main text. That is, the model for the treatment effect was specified as a linear function of the moderator, namely $\beta_{0}Z_{t}+\beta_{1}Z_{t}M_{t}=(\beta_{0}+\beta_{1}M_{t})Z_{t}$. To assess the sensitivity of linearity assumption,we explore potential non-linearity in the interaction term between the causal effect of competition and additional weeks in the study, namely $f(M_{t})Z_{t}$, where $f$ is a smooth nonlinear function. Here we fit $f$ using penalized basis spline by *gam* function from *mgcv* R package and natural cubic spline from *ns* function in R. The penalized basis spline models were fit using restricted maximum likelihood method and (REML) and thin plate regression spline as smoothing basis.

Third, to assess the sensitivity of our results to different missingness patterns, the paper’s results, which were from multiple imputation analysis, were compared with complete-case analysis after introducing a certain missingness pattern. Here we examined two different missingness patterns: dropout and weekly missingness. For dropout complete-case analysis, we removed imputed data from interns who dropped out from the study early. For example, if a user does not have any data points after Sep 1, 2020, all the imputed data points for this user after Sep 1, 2020, were removed. For weekly missingness complete-case analysis, we removed weeks with a large percentage of missing data in the outcome of interest. For example, we eliminated all weeks where more than 5 data points were missing before performing complete-case analysis.

**eResults**

**Mood-score related analyses**

The parameter estimates for linear models assessing the marginal and time-varying causal effect of competition on weekly average daily mood score were shown in eTable 6. From the marginal-effect model, we concluded that on average competition tended to improve the daily mood score with an estimated effect of 0.02 (SE 0.02, p=0.35). We also concluded that the additional weeks in the study was a significantly negative moderator of the causal effect of competition on daily mood score with an estimated moderation of -0.01 (SE 0.01, p=0.05) from the time-varying-effect model. The time-varying-effect plot showed that being in the competition arm had a significant positive effect on daily mood score at the early stage of the study and the effect waned over time.

The parameter estimates for linear models assessing the marginal and time-varying causal effect of competition on participation rate of daily mood survey were shown in eTable 5. Also, the estimated causal effect of competition on participation rate of daily mood survey at different weeks was shown in eFigure 4. We concluded that the competition did not affect the participation rate of daily mood surveys marginally. From eFigure 4, we can observe an interesting fact that the competition decreased the intern's participation rate of self-report mood surveys early in the study. Other than push notifications including life insight and tips received by all interns, the ones assigned to the competition arm received additional competition-related messages (Table 1) four times per week, which might make those less responsive to the push notifications and more possible to ignore the mood survey completion reminder 8:00 pm every night. Daily step and survey data were collected objectively through the fitness tracker, which was less sensitive to push notification fatigue. Intensive mHealth push notifications (overtreatment) may lead to inferior treatment effect; therefore, this gives rise to the need for just-in-time adaptive intervention (JITAI), which can deliver mHealth intervention optimally.

**Sensitivity analysis**

***Sensitivity of complete-case analysis***

For primary aim, the estimate of the marginal causal effect of competition on step count from multiple imputation analysis was 111.5 (SE 40.4, p=0.01) steps, compared to 102.6 (SE 46.9, p=0.03) steps from complete-case analysis. The estimate of the causal effect of competition on sleep duration from multiple imputation analysis was -0.7 (SE 1.8, p=0.69) minutes, compared to -0.2 (SE 2.0, p=0.93) minutes from complete-case analysis. We can conclude that the conclusions for primary aim were mildly sensitive to missingness mechanisms.

For secondary aims, the estimate of the moderation of additional weeks in the study on the causal effect of competition on step count was -9.1 (SE 11.6, p=0.43) steps/week from multiple imputation analysis, compared to -6.9 (SE 11.7, p=0.55) steps/week from complete-case analysis. The estimate of the moderation of additional weeks in the study on the causal effect of competition on sleep duration was -1.9 (SE 0.6, p=0.003) minutes/week from multiple imputation analysis, compared to -1.1 (SE 0.6, p=0.06) minutes/week from complete-case analysis. The estimate of the moderation of competing within the same institution or specialty on the causal effect of competition on step count was -114.9 (SE 93.7, p=0.22) steps and 26.1(SE 74.7, p=0.73) steps from multiple imputation analysis, compared to -172.7 (SE 104.1, p=0.10) steps and 71.2 (SE 77.7, p=0.36) steps from complete-case analysis. The estimate of the moderation of competing within the same institution or specialty on the causal effect of competition on sleep duration was 0.4 (SE 3.1, p=0.90) minutes and -1.9 (SE 3.2, p=0.57) minutes from multiple imputation analysis, compared to -1.7 (SE 4.1, p=0.68) minutes and 2.1 (SE 4.4, p=0.63) minutes from complete-case analysis. We can conclude that the conclusions for moderation of additional weeks in the study on causal effect of competition were insensitive to missingness mechanisms, while the conclusions for moderation of competing within the same institution or specialty on causal effect of competition were sensitive to missingness mechanisms. The size of the estimated moderation of competing within the same institution was enlarged when performing complete-case analysis and the sign of the moderation remained negative, matching the conclusions made in the main text.

For mood-related analysis, the estimate of the marginal causal effect of competition on mood score from multiple imputation analysis was 0.02 (SE 0.02,p=0.35), compared to 0.01 (SE 0.02, p=0.32) from complete-case analysis. The estimate of the moderation of additional weeks in the study on the causal effect of competition on mood score was -0.01 (SE 0.01, p=0.05) from multiple imputation analysis, compared to -0.01 (SE 0.00, p=0.05) from complete-case analysis. We can conclude that the conclusions for mood-related analysis were insensitive to missingness mechanisms.

The estimates of all the models mentioned above can be obtained through eFigure 2-4,6.

***Sensitivity of non-linear moderation of treatment effect***

The estimated causal effect of competition on step count or sleep duration at different weeks from nonlinear regression was plotted in eFigure 5. From the plots, we can see that linearity assumption is appropriate for our analysis.

The estimated causal effect of competition on participation rate of step count, sleep duration and mood survey at different weeks from nonlinear regression was plotted in eFigure 6. From the plots, we can notice some evidence of non-linearity, while linearity assumption is still appropriate for easy interpretation.

***Sensitivity of missingness patterns***

For primary aim, the estimate of marginal causal effect of competition on step count was 111.5 (SE 40.4) steps from multiple imputation analysis, compared to 93.1 (SE 45.7) steps from complete-case analysis with dropout and 91.1 (SE 45.7) steps from complete-case analysis with weekly missingness. The estimate of marginal causal effect of competition on sleep duration was -0.7(SE 1.8) minutes from multiple imputation analysis, compared to -0.5 (SE 1.7) steps from complete-case analysis with dropout and -0.5 (SE 1.8) minutes from complete-case analysis with weekly missingness. We concluded that the conclusions of the primary aim were robust to both dropout and weekly data missingness.

For secondary aims, the estimate of the moderation of additional weeks in the study on the causal effect of competition on step count was -9.1 (SE 11.6) steps/week from multiple imputation analysis, compared to -7.7 (SE 11.7) steps/week from complete-case analysis with dropout and -2.6 (SE 11.6) steps/week from complete-case analysis with weekly missingness. The estimate of the moderation of additional weeks in the study on the causal effect of competition on sleep duration was -1.9 (SE 0.6) minutes/week from multiple imputation analysis, compared to -1.6 (SE 0.6) minutes/week from complete-case analysis with dropout and -1.3 (SE 0.6) minutes/week from complete-case analysis with weekly missingness. The estimate of the moderation of competing within the same institution or specialty on the causal effect of competition on step count was -114.9 (SE 93.7) steps and 26.1 (SE 74.7) steps from multiple imputation analysis, compared to -171.4 (SE 102.1) steps and 25.4 (SE 77.9) steps from complete-case analysis with dropout and -186.2 (SE 105.9) steps and 27.1 (SE 80.8) steps from complete-case analysis with weekly missingness. The estimate of the moderation of competing within the same institution or specialty on the causal effect of competition on sleep duration was 0.4 (SE 3.1) minutes and -1.9 (SE 0.6) minutes from multiple imputation analysis, compared to -2.0 (SE 3.9) minutes and -1.1 (SE 4.1) minutes from complete-case analysis with dropout and -1.2 (SE 4.2) minutes and 0.4 (SE 4.5) minutes from complete-case analysis with dropout. We can conclude that the conclusions for the effect of two moderators on the causal effect of competition were insensitive to dropout and weekly missingness, except that the moderation intra-institution competition is sensitive to the weekly missingness and dropout. The sign the moderation remained negative; however, the effect sizes were increased.

For mood-related analysis, the estimate of marginal causal effect of competition on mood score was 0.02 (SE 0.02) from multiple imputation analysis, compared to 0.03 (SE 0.02) from complete-case analysis with dropout and 0.02 (SE 0.02) from complete-case analysis with weekly missingness. The estimate of moderation of additional weeks in the study on causal effect of competition on mood score was -0.01 (SE 0.01) from multiple imputation analysis, compared to -0.02 (SE 0.01) steps from complete-case analysis with dropout and -0.02 (SE 0.01) from complete-case analysis with weekly missingness. We can conclude that the conclusions of mood-related analysis were insensitive to dropout and weekly missingness.

The estimates of all the models mentioned above can be obtained through eTable 7-10.

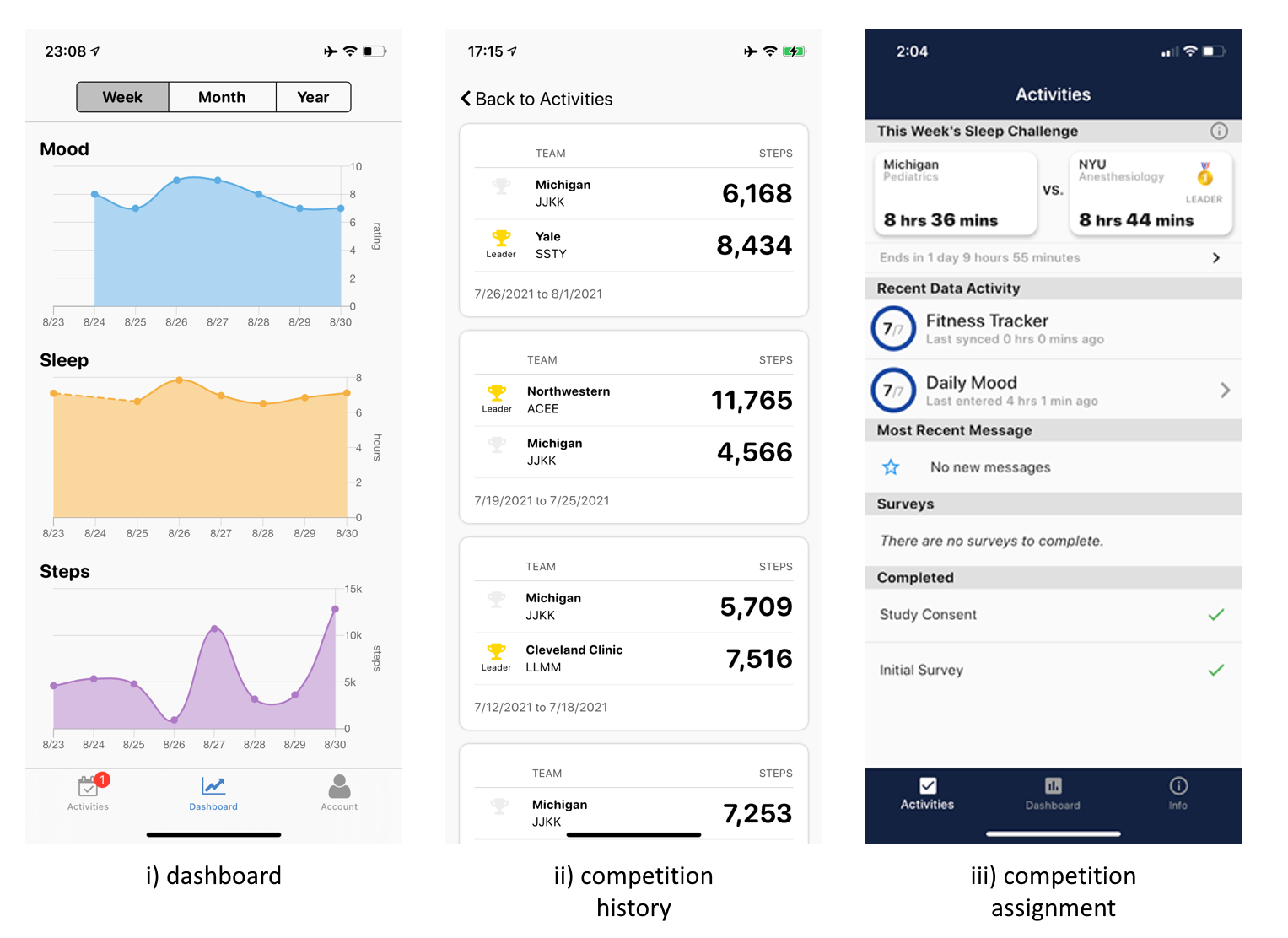

**eFigure 1: Screenshots of the app dashboard, previous competition history and weekly competition assignment. The screenshot of previous competition history contains pseudo program names.**

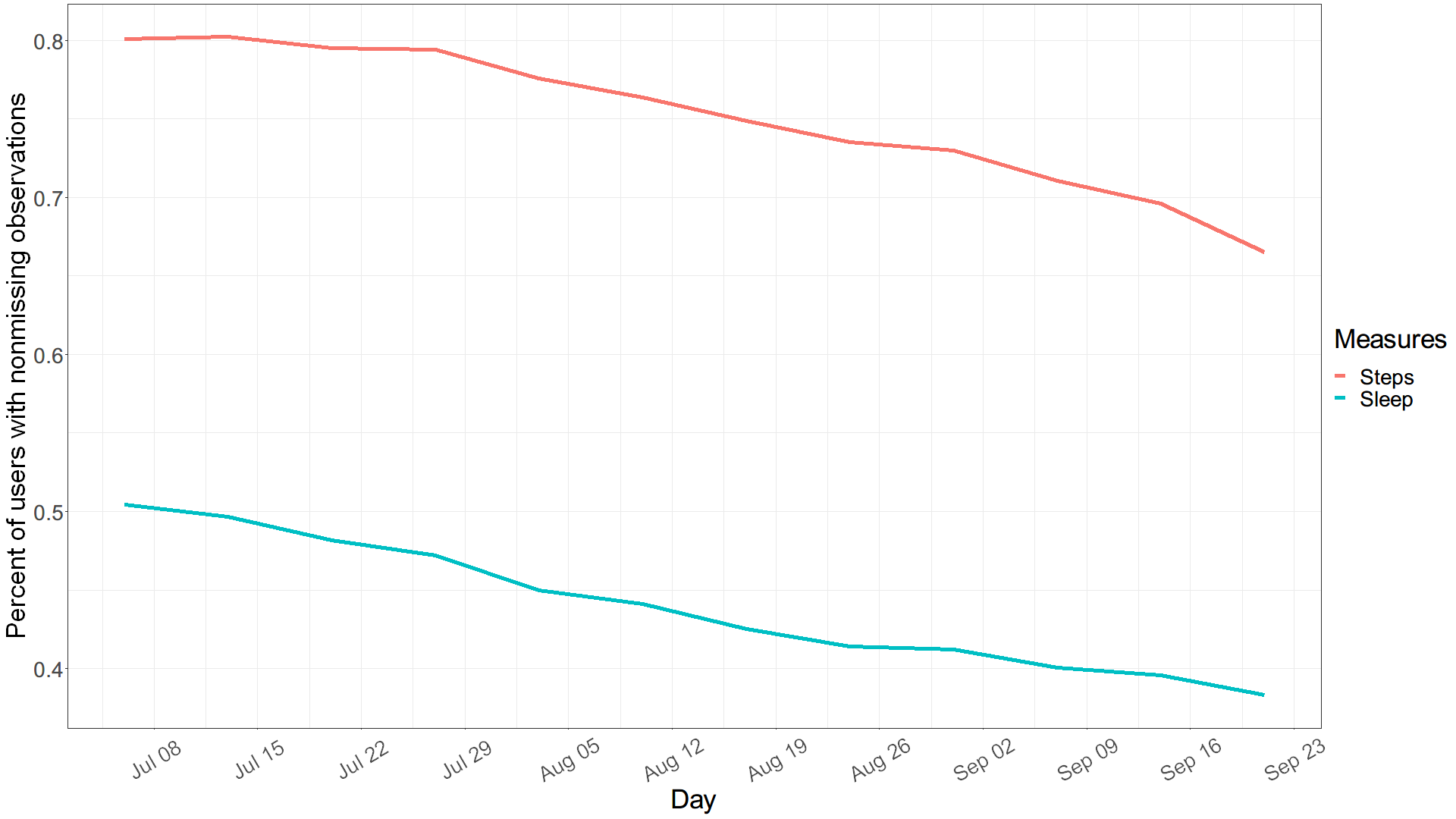
**eFigure 2: Percentage of interns with nonmissing step and sleep observation for each day in the study**

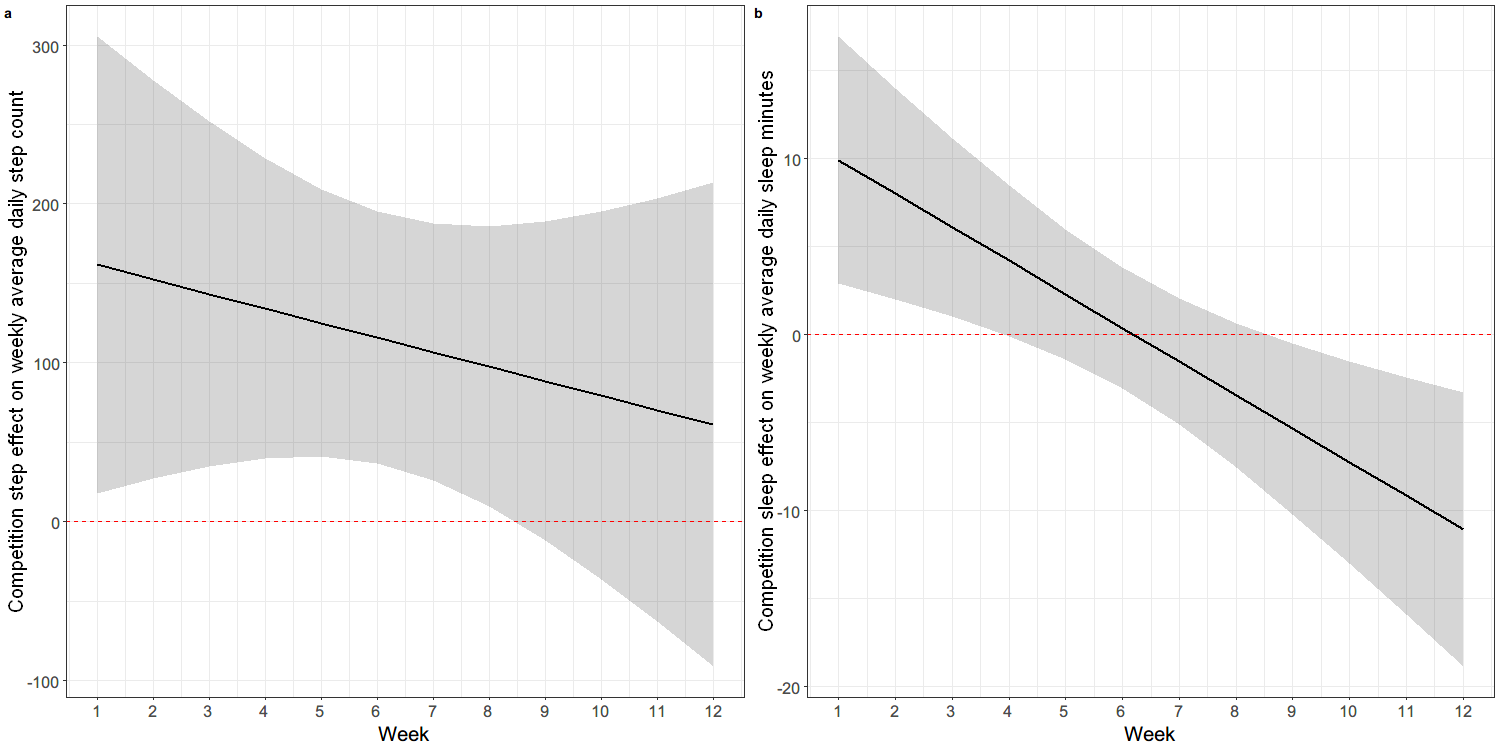
**eFigure 3: a) Estimated causal effect of competition step on weekly average daily step count at different weeks. b) Estimated causal effect of competition sleep on weekly average daily sleep minutes at different weeks. Shaded area indicates 95% CI. Red dotted line indicates no effect.**

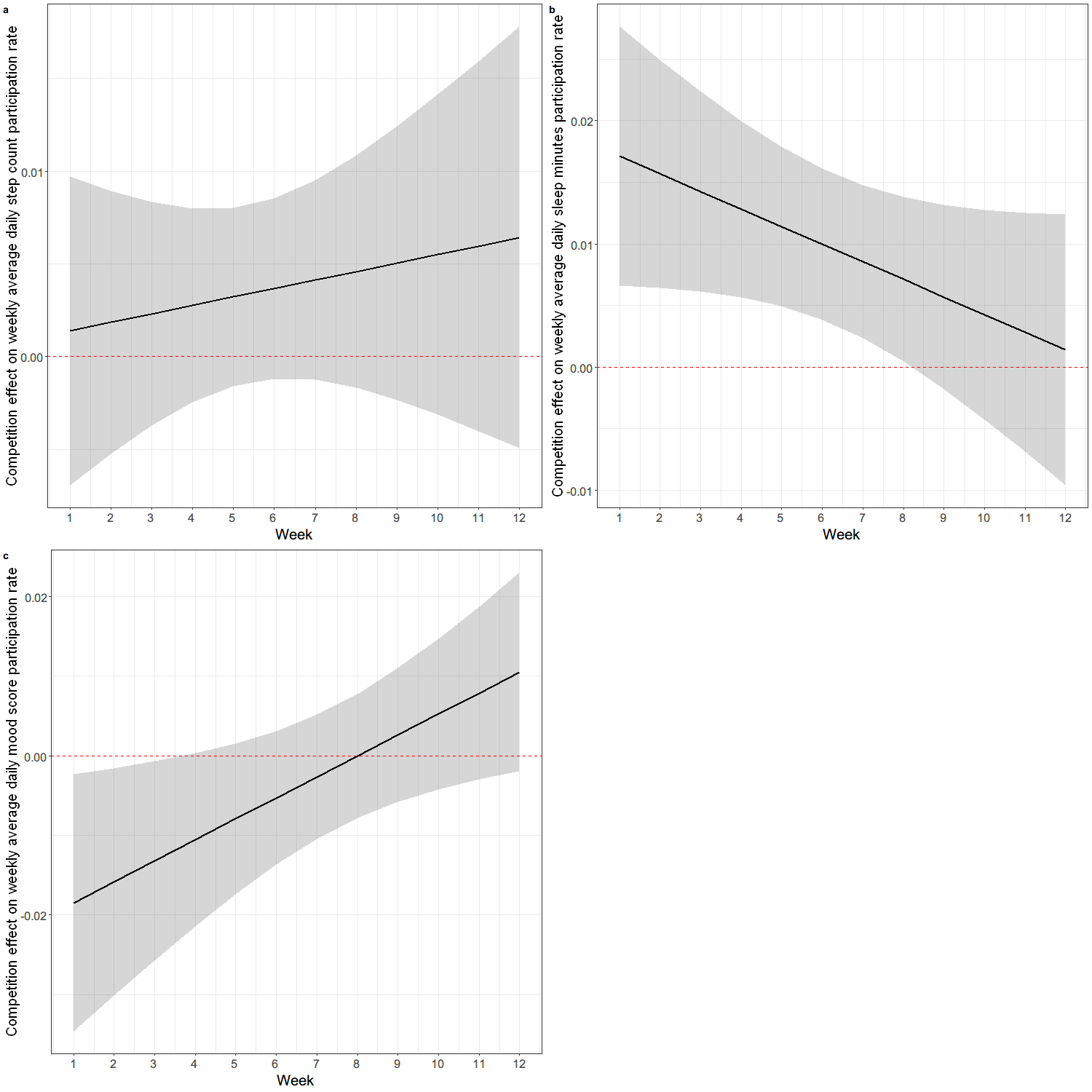
**eFigure 4: Estimated causal effect of competition on the participation rate of a) daily step count, b) daily sleep minutes c) daily mood survey, at different weeks. Shaded area indicates 95% CI. Red dotted line indicates the effect being 0.**

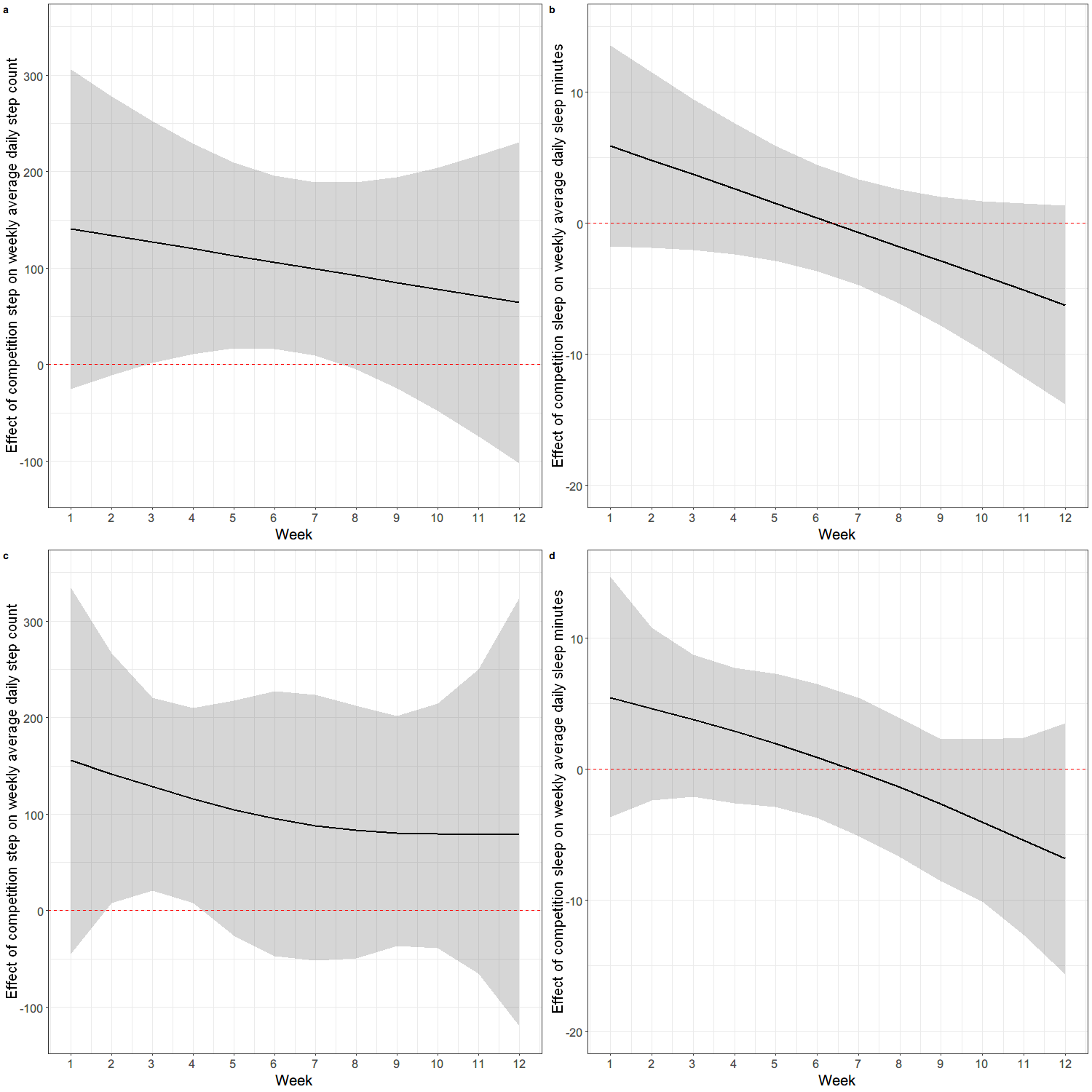
**eFigure 5: a,b)Estimated causal effect of competition on weekly average daily a) step count and b) sleep duration at different weeks fitted using penalized basis spline, and shaded area is 95% CI. c,d) Estimated causal effect of competition on weekly average daily c) step count and d) sleep duration at different weeks fitted using natural cubic spline, and shaded area indicates 95% CI. Red dotted line indicates no effect.**

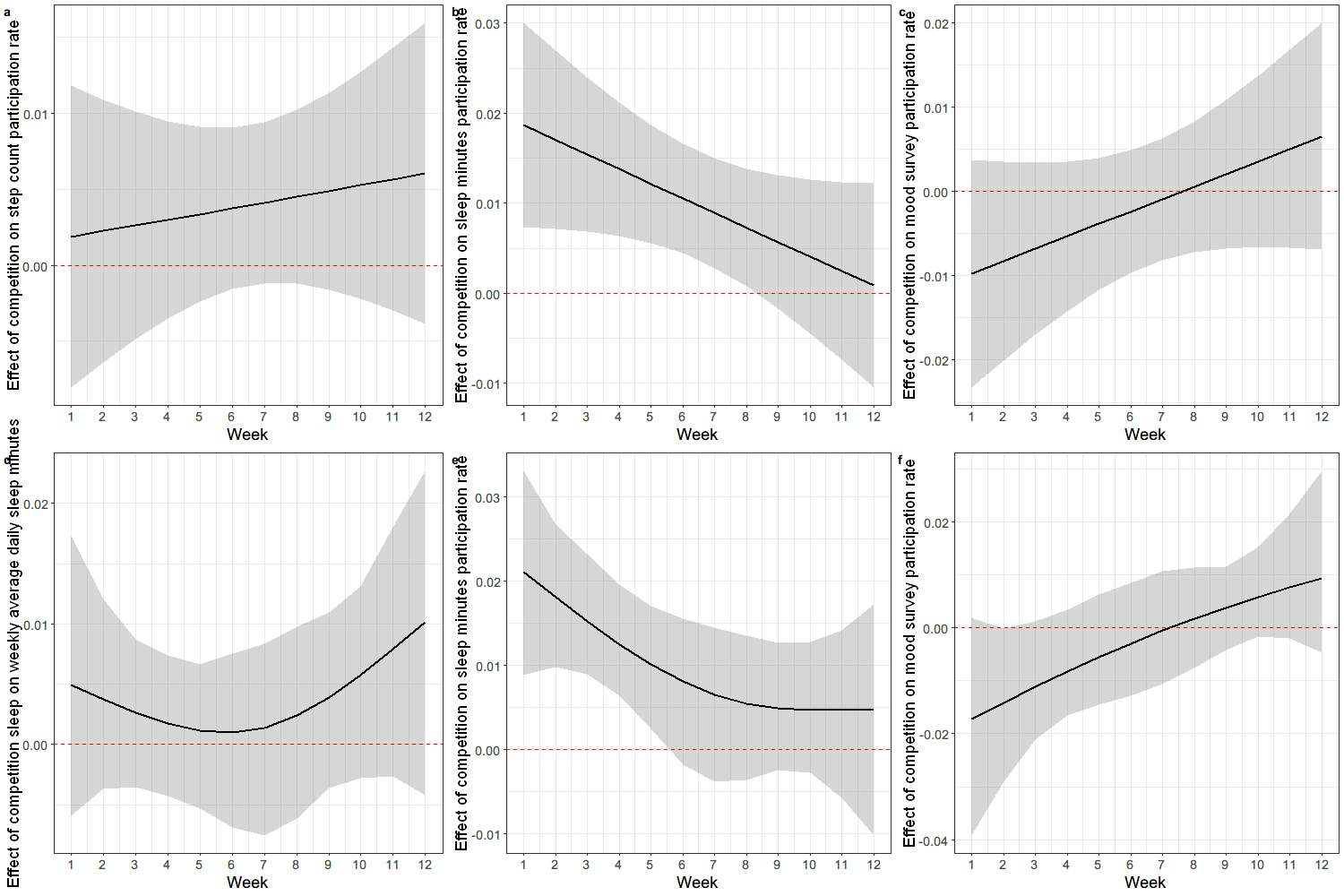
**eFigure 6 : a,b,c) Estimated causal effect of competition on participation rate of a) step count, b) sleep minutes, c) mood score at different weeks using penalized basis spline, and shaded area is 95% CI. d,e,f) Estimated causal effect of competition on participation rate of d) step count, e) sleep minutes, f) mood score at different weeks using natural cubic spline, and shaded area indicates 95% CI. Red dotted line indicates no effect.**

**eTable 1. Examples of different types of push notifications**

| **Message types** | **Time** | **Examples** |
| --- | --- | --- |
| Alert of competition types and opponent | Sunday 9:00 pm | MGH Surgery faces off against Northwestern Internal Medicine in this week’s step competition! |
| Competition score status update | Wednesday 9:00 pm and Saturday 11:00 am | Yale Psychiatry is leading in this week’s sleep challenge with an average of 8 hrs 41 min. Let’s see who will win! |
| Competition final result | Monday 12:00 pm | Michigan Pediatrics comes out on top of this week’s step challenge against NYU Anesthesiology. Great job to both teams! |

**eTable 2: Parameter estimates for linear model using complete-case and 20-time multiple imputation dataset, assessing marginal causal effect of competition on daily step count and sleep duration**

| **Outcome & Competition type** |  | **Complete case** | | **Multiple Imputation** | |
| --- | --- | --- | --- | --- | --- |
|  | **Parameter** | **Estimate** | **95% CI** | **Estimate** | **95% CI** |
| **Step** | Intercept | 7779.9 | 7635.3,7924.4 | 7679.3 | 7552.0,7806.6 |
|  | Week | -12.6 | -23.0,-2.2 | -18.0 | -28.6,-7.4 |
|  | Competition Step | 102.6 | 10.8,194.4 | 111.5 | 32.2,190.8 |
| **Sleep** | Intercept | 415.2 | 409.2,421.1 | 416.6 | 411.8,421.5 |
|  | Week | 0.3 | -0.1,0.8 | 0.0 | -0.3,0.3 |
|  | Competition Sleep | -0.2 | -4.2,3.8 | -0.7 | -4.3,2.9 |

**eTable 3: Parameter estimates for linear model using complete-case and 20-time multiple imputation dataset, assessing time-varying causal effect of competition on daily step count and sleep duration**

| **Outcome & Competition type** |  | **Complete case** | | **Multiple Imputation** | |
| --- | --- | --- | --- | --- | --- |
|  | **Parameter** | **Estimate** | **95% CI** | **Estimate** | **95% CI** |
| **Step** | Intercept | 7768.6 | 7617.4,7919.8 | 7664.5 | 7529.2,7799.8 |
|  | Week | -10.4 | -24.4,3.6 | -15.1 | -28.8,-1.4 |
|  | Competition Step | 140.6 | 0.9,280.4 | 161.5 | 17.2,305.8 |
|  | Week: Competition Step | -6.9 | -29.8,15.9 | -9.1 | -32.0,13.8 |
| **Sleep** | Intercept | 413.4 | 407.2,419.6 | 413.5 | 408.3,418.8 |
|  | Week | 0.6 | 0.1,1.2 | 0.5 | 0.1,1.0 |
|  | Competition Sleep | 5.9 | -0.8,12.6 | 9.9 | 2,7,17.1 |
|  | Week: Competition Sleep | -1.1 | -2.2,0.0 | -1.9 | -3.1,-0.7 |

**eTable 4: Parameter estimates for linear models using complete-case and 20-time multiple imputation dataset, assessing moderation of competing within the same institution on causal effect of competition on step count and sleep duration**

| **Outcome & Competition type** |  | **Complete Case** | | **Multiple Imputation** | |
| --- | --- | --- | --- | --- | --- |
|  | **Parameter** | **Estimate** | **95% CI** | **Estimate** | **95% CI** |
| **Step** | Intercept | 7766.8 | 7614.0,7919.6 | 7664.5 | 7527.6,7801.4 |
|  | Week | -10.4 | -24.4,3.6 | -15.1 | -28.8,-1.4 |
|  | Competition Step | 167.2 | 21.6,312.8 | 182.1 | 31.7,332.5 |
|  | Week: Competition Step | -11.4 | -34.9,12.2 | -11.3 | -34.9,12.2 |
|  | Competition Step : Same Institution Competition | -172.7 | -376.7,31.3 | -114.9 | -299.3,69.6 |
|  | Competition Step : Same Specialty Competition | 71.2 | -81.2,223.5 | 26.1 | -120.9,173.2 |
| **Sleep** | Intercept | 413.4 | 407.3,419.6 | 413.5 | 408.3,418.8 |
|  | Week | 0.6 | 0.1,1.2 | 0.5 | 0.1,1.0 |
|  | Competition Sleep | 5.9 | -1.4,13.3 | 10.1 | 2.8,17.5 |
|  | Week: Competition Sleep | -1.1 | -2.3,0.0 | -1.9 | -3.1,-0.6 |
|  | Competition Sleep : Same Institution Competition | -1.7 | -9.6,6.3 | 0.4 | -5.8,6.6 |
|  | Competition Sleep : Same Specialty Competition | 2.1 | -6.5,10.8 | -1.9 | -8.3,4.6 |

**eTable 5: Parameter estimates for linear models, assessing marginal and time-varying causal effect of competition on participation rate of daily step count, sleep duration and mood score(*100)**

| **Model** |  | **Step** | | **Sleep** | | **Mood** | |
| --- | --- | --- | --- | --- | --- | --- | --- |
|  | **Parameter** | **Estimate** | **95% CI** | **Estimate** | **95% CI** | **Estimate** | **95% CI** |
| **Main-effect** | Intercept | 76.8 | 76.0,77.5 | 43.5 | 42.8,44.3 | 51.2 | 49.7,52.6 |
|  | Week | -0.4 | -0.4,-0.3 | -0.2 | -0.3,-0.1 | -0.6 | -0.7,-0.5 |
|  | Competition | 0.4 | -0.1,0.9 | 0.9 | 0.3,1.5 | -0.4 | -1.2,0.4 |
| **Time-varying-effect** | Intercept | 76.9 | 76.1,77.7 | 43.2 | 42.3,44.1 | 51.8 | 50.3,53.4 |
|  | Week | -0.4 | -0.5,-0.3 | -0.1 | -0.2,0.0 | -0.8 | -0.9,-0.6 |
|  | Competition | 0.1 | -0.7,1.0 | 1.7 | 0.7,2.8 | -1.8 | -3.5,-0.2 |
|  | Week: Competition | 0.1 | -0.1,0.2 | -0.1 | -0.3,0.0 | 0.3 | 0.0,0.5 |

**eTable 6: Parameter estimates for linear model using complete-case and 20-time multiple imputation dataset, assessing marginal and time-varying causal effect of competition on mood score(*100)**

| **Outcome & Competition type** | **Model** |  | **Complete case** | | **Multiple Imputation** | |
| --- | --- | --- | --- | --- | --- | --- |
|  |  | **Parameter** | **Estimate** | **95% CI** | **Estimate** | **95% CI** |
| **Mood** | **Main-effect model** | Intercept | 720.1 | 714.8,725.5 | 736.3 | 723.4,749.2 |
|  |  | Week | -0.2 | -0.6,0.2 | -1.7 | -2.3,-1.0 |
|  |  | Competition | -1.1 | -4.2,2.0 | 1.9 | -2.1,6.0 |
|  | **Time-varying-effect model** | Intercept | 718.0 | 712.3,723.8 | 733.4 | 720.1,746.7 |
|  |  | Week | 0.2 | -0.4,0.8 | -1.1 | -1.9,-0.3 |
|  |  | Competition | 3.6 | -1.7,8.9 | 8.3 | 1.3,15.3 |
|  |  | Week: Competition | -0.9 | -1.7,0.0 | -1.2 | -2.3,0.0 |

**eTable 7: Sensitivity analyses for assessing different missing patterns on marginal causal effect of competition on daily step count and sleep duration**

| **Missing pattern** | **Outcome & Competition type** | **Parameter** | **Estimate** | **95% CI** |
| --- | --- | --- | --- | --- |
| **Dropout** | **Step** | Intercept | 7735.4 | 7604.7, 7866.1 |
|  |  | Week | -14.4 | -25.0, -3.7 |
|  |  | Competition | 93.1 | 3.5, 182.8 |
|  | **Sleep** | Intercept | 416.1 | 411.4, 420.8 |
|  |  | Week | 0.1 | -0.4, 0.5 |
|  |  | Competition | -0.5 | -3.9, 2.8 |
| **Weekly missingness** | **Step** | Intercept | 7796.7 | 7672.9, 7920.5 |
|  |  | Week | -15.2 | -26.0, -4.3 |
|  |  | Competition | 91.1 | 1.4,180.7 |
|  | **Sleep** | Intercept | 416.2 | 411.8, 420.5 |
|  |  | Week | 0.4 | -0.1,0.8 |
|  |  | Competition | -0.6 | -4.1, 3.0 |

**eTable 8: Sensitivity analyses for** **assessing different missing patterns on time-varying causal effect of competition on daily step count and sleep duration**

| **Missing pattern** | **Outcome & Competition type** | **Parameter** | **Estimate** | **95% CI** |
| --- | --- | --- | --- | --- |
| **Dropout** | **Step** | Intercept | 7722.9 | 7589.9, 7856.0 |
|  |  | Week | -11.9 | -25.8, 1.9 |
|  |  | Competition | 135.3 | -7.0, 277.5 |
|  |  | Week: Competition | -7.7 | -30.6, 15.2 |
|  | **Sleep** | Intercept | 413.7 | 408.3, 419.0 |
|  |  | Week | 0.5 | -0.0,1.1 |
|  |  | Competition | 8.1 | 1.1, 15.1 |
|  |  | Week: Competition | -1.6 | -2.8, -0.4 |
| **Weekly missingness** | **Step** | Intercept | 7792.4 | 7664.9, 7919.8 |
|  |  | Week | -14.4 | -28.6, -0.2 |
|  |  | Competition | 105.5 | -35.9, 246.9 |
|  |  | Week: Competition | -2.6 | -25.5, 20.2 |
|  | **Sleep** | Intercept | 414.2 | 409.4, 419.0 |
|  |  | Week | 0.7 | 0.2, 1.3 |
|  |  | Competition | 6.4 | -0.2, 12.9 |
|  |  | Week: Competition | -1.3 | -2.4, -0.1 |

**eTable 9: Sensitivity analyses for assessing different missing patterns on moderation of competing within the same institution or specialty on causal effect of competition on daily step count and sleep duration**

| **Missing pattern** | **Outcome & Competition type** | **Parameter** | **Estimate** | **95% CI** |
| --- | --- | --- | --- | --- |
| **Dropout** | **Step** | Intercept | 7722.8 | 7588.2, 7857.4 |
|  |  | Week | -12.0 | -25.8, 1.9 |
|  |  | Competition Step | 167.7 | 20.8, 314.6 |
|  |  | Week: Competition Step | -10.5 | -33.9, 12.9 |
|  |  | Competition Step : Same Institution Competition | -171.4 | -371.9, 29.0 |
|  |  | Competition Step : Same Specialty Competition | 25.4 | -127.6, 178.3 |
|  | **Sleep** | Intercept | 413.7 | 408.4, 419.0 |
|  |  | Week | 0.5 | -0.0, 1.1 |
|  |  | Competition Sleep | 8.7 | 1.4, 16.1 |
|  |  | Week: Competition Sleep | -1.6 | -2.8, -0.4 |
|  |  | Competition Sleep : Same Institution Competition | -2.0 | -9.7, 5.7 |
|  |  | Competition Sleep : Same Specialty Competition | -1.1 | -9.1, 6.9 |
| **Weekly missingness** | **Step** | Intercept | 7791.9 | 7663.1,7920.7 |
|  |  | Week | -14.4 | -28.6,-0.2 |
|  |  | Competition Step | 140.8 | -6.6, 288.3 |
|  |  | Week: Competition Step | -5.7 | -29.1, 17.7 |
|  |  | Competition Step: Same Institution Competition | -186.2 | -394.1, 21.6 |
|  |  | Competition Step: Same Specialty Competition | 27.1 | -131.4, 185.6 |
|  | **Sleep** | Intercept | 414.2 | 409.4, 419.0 |
|  |  | Week | 0.7 | 0.2, 1.3 |
|  |  | Competition Sleep | 6.6 | -0.5, 13.6 |
|  |  | Competition Sleep : Same Institution Competition | -1.2 | -9.5, 7.0 |
|  |  | Competition Sleep : Same Specialty Competition | 0.4 | -8.4, 9.2 |

**eTable 10: Sensitivity analyses for assessing different missing patterns on time-varying causal effect of competition on causal effect of competition on daily mood score (*100)**

| **Missing pattern** | **Model** | **Parameter** | **Estimate** | **95% CI** |
| --- | --- | --- | --- | --- |
| **Dropout** | **Main-effect model** | Intercept | 730.3 | 717.2, 743.4 |
|  |  | Week | -1.7 | -2.4, -0.9 |
|  |  | Competition | 2.7 | -2.0, 7.4 |
|  | **Time-varying-effect model** | Intercept | 725.8 | 712.3, 739.3 |
|  |  | Week | -0.8 | -1.8, 0.2 |
|  |  | Competition | 12.6 | 4.1, 21.1 |
|  |  | Week: Competition | -1.8 | -3.2, -0.4 |
| **Weekly missingness** | **Main-effect model** | Intercept | 739.3 | 724.2, 754.4 |
|  |  | Week | -1.7 | -2.4, -1.1 |
|  |  | Competition | 2.1 | -2.7, 6.9 |
|  | **Time-varying-effect model** | Intercept | 734.6 | 719.2,750.0 |
|  |  | Week | -0.9 | -1.7, -0.0 |
|  |  | Competition | 12.2 | 0.4,20.2 |
|  |  | Week: Competition | -1.8 | -3.1, -0.6 |
